# Mortality of Diabetes-related Acute Metabolic Emergencies in COVID-19 patients: a systematic review and meta-analysis

**DOI:** 10.1101/2021.01.12.21249697

**Authors:** Vasileios P. Papadopoulos, Peny Avramidou, Stefania-Aspasia Bakola, Dimitra-Geogia Zikoudi, Ntilara Touzlatzi, Marios-Vasileios Koutroulos, Dimitrios K. Filippou

**Affiliations:** ENARGEIA Medical Ltd, Xanthi, Greece; Department of Internal Medicine, Xanthi General Hospital, Xanthi, Greece; Laboratory of Anatomy, Medical School, National and Kapodistrian University of Athens, Athens, Greece

**Author notes:** Corresponding author: Vasileios P. Papadopoulos, 6, Elpidos str., GR-67131 Xanthi, Greece. Conflicts of interest: None declared.

**Keywords:** COVID-19, diabetic ketoacidosis, hyperglycaemic hyperosmolar state, euglycaemic diabetic ketoacidosis, meta-analysis

## Abstract

**Purpose:** Little is known on the mortality rate in COVID-19 related acute metabolic emergencies, namely diabetic ketoacidosis (DKA), hyperglycaemic hyperosmolar state (HHS), combined DKA/HHS, and euglycaemic diabetic ketoacidosis (EDKA).

**Methods:** A systematic literature review was conducted using EMBASE, PubMed/Medline, and Google Scholar from January 1, 2020 to January 9, 2021 to identify all case report series, cross-sectional studies, and meta-analyses of case reports describing mortality rate in DKA, HHS, and EDKA, in COVID-19 patients. The Joanna Briggs Institute critical appraisal checklist for case reports was used for quality assessment.

**Results:** From 313 identified publications, 4 fulfilled the inclusion criteria and analyzed qualitatively and quantitatively. A systematic review and meta-analysis with subgroup analyses examined mortality rate in a total of 152 COVID-19 patients who had developed DKA, HHS, combined DKA/HHS, or EDKA. Combined mortality rate and confidence intervals (CI) were estimated using random effects model. The study was registered to PROSPERO database (ID: 230737).

**Results:** Combined mortality rate was found to be 27.1% [95% CI: 11.2-46.9%]. Heterogeneity was considerable (I^2^=83%; 95% CI: 56-93%), corrected to 67% according to Von Hippel adjustment for small meta-analyses. Funnel plot presented no apparent asymmetry; Egger’s and Begg’s test yielded in P=0.44 and P=0.50, respectively. Sensitivity analysis failed to explain heterogeneity.

**Conclusion:** COVID-19 related acute metabolic emergencies (DKA, HHS, and EDKA) are characterized by considerable mortality; thus, clinicians should be aware of timely detection and immediate treatment commencing.

## Introduction

Diabetes mellitus (DM) has been recognized as a major risk factor for unfavorable outcomes in patients with COVID-19 caused by severe acute respiratory syndrome coronavirus 2 (SARS-CoV-2) [1]. The underlying pathophysiology of COVID-19 and DM intertwining has not been totally explained; however, diabetes patients have an increased risk of infection and acute respiratory distress syndrome compared with the general population [2] [3] [4] [5] [6], while direct cytopathic effects of SARS-CoV-2 on pancreatic b-cell populations have been proposed [7].

COVID-19 is associated with hyperglycaemic emergencies as diabetic ketoacidosis (DKA), hyperglycaemic hyperosmolar State (HHS), euglycaemic diabetic ketoacidosis (EDKA), and combined DKA/HHS [8] [9]. The over-activity of immune system might further explain COVID-19-related severe and resistant to conventional therapy DKA episodes [10]. High mortality in COVID-19 and diabetic ketoacidosis has been reported in a single letter [11]. In contrast, two small case series exhibited significantly lower mortality rates that range from 7.7% [8] to 12.9% [12]. Additionally, a few dozen of case reports concerning acute emergencies related to glucose metabolism in COVID-19 patients have been reported, all reviewed in a very recent meta-analysis [13]. The present study aimed to provide further evidence regarding the mortality rate in COVID-19-related acute metabolic emergencies (DKA, HHS, combined DKA/HHS, and EDKA) by identifying all relevant studies and summarize their results.

## Materials and Methods

### Literature search

A systematic literature review was conducted using PubMed/MEDLINE and EMBASE databases from January 01, 2020 until January 09, 2021 to identify all case report series, cross-sectional studies and meta-analyses of case reports written in English and describing mortality rate in DKA, HHS, combined DKA/HHS, and EDKA in COVID-19 patients. The Google Scholar and ResearchGate databases were used as an additional pool of published data, dissertations and other unpublished work; an iterative search was performed until no additional publication could be traced. Personal communication was followed where needed. The relevant protocol was registered in PROSPERO database on January 10, 2021 (ID: 230737).

### Study selection

The review was conducted using a search strategy that included the PubMed search terms [diabetes] AND [ketoacidosis] AND [covid] OR [diabetic] AND [ketoacidosis] AND [covid] OR [euglycemic] AND [diabetic] AND [ketoacidosis] AND [covid] OR [hyperglycaemic] AND [hyperosmolar] AND [state] AND [covid].

Eligible studies were all that (1) are written in English; (2) are either report series or cross-sectional-studies or meta-analyses of case reports; (3) report mortality rate in COVID-19 patients who had developed either DKA, or HHS, or EDKA, or combined DKA/HHS or enough data to compute it; (4) report a relevant measure of statistical significance; (5) are not duplicates.

Six reviewers (V.P., M.-V.K., N.T., S.-A.B., P.A., D.-G.-Z.) screened the total number of initially identified studies, applied eligibility criteria and selected studies for inclusion in the systematic review working simultaneously as three independent couples of investigators (one for screening and the other for checking decisions). These three couples were blinded to each other’s decisions. D.F. was responsible to dissolve any disagreement. No specific software was used for recording decisions; all data were transformed to a suitable Word table. Sources of financial support were traced where possible.

### Outcome measures

The present study was conducted in accordance to the MOOSE reporting guidelines for observational studies [14]. Mortality rates were compared between different types of acute metabolic emergencies in diabetes (DKA, HHS, EDKA, and DKA/HHS) was performed. AMSTAR 2 checklist was used to confirm the high quality of the present meta-analysis [15].

### Data extraction

A structured data collection was used to extract the following data from each eligible study: title of the study, name of the first author, year of publication, study design, country where the study was conducted, total number of patients per type of acute metabolic emergency, total number of survivors, and total number of non-survivors. The data extraction process was carried out by six reviewers (V.P., M.-V.K., N.T., S.-A.B., P.A., D.-G.-Z.) who performed data extraction working simultaneously as three independent couples of investigators (consisting of one for extracting data and another for checking the extracted data); the process performed manually and the three couples were blinded to each other’s decisions. D.F. closely observed the process and was responsible for any discordance.

### Quality assessment of the studies

The Joanna Briggs Institute (JBI) critical appraisal checklist for case reports, which includes 8 questions addressing the internal validity and risk of bias of case reports designs, particularly confounding and information bias, in addition to the importance of clear reporting, was used for quality assessment of case series [17]. All studies that failed to fulfill requirements of first six questions were considered as of “suboptimal quality”; controversially, an “optimal quality” remark was given.

Moreover, the JBI critical appraisal list for case control studies, which includes 12 questions addressing the internal validity and risk of bias of case control studies, was used for quality assessment of case series [16] [17]. All studies that were characterized as of “fair” or “poor” quality were considered as of “suboptimal quality”; controversially, an “optimal quality” remark was given.

Furthermore, quality of evidence was approached using GRADE (Grading of Recommendations, Assessment, Development and Evaluations), transparent framework for developing and presenting summaries of evidence [18-20]. GRADE level of evidence was rated down for risk of bias, imprecision, inconsistency, indirectness, and publication bias, whereas was rated up for large magnitude of effect.

The process was carried out by six reviewers (V.P., M.-V.K., N.T., S.-A.B., P.A., D.-G.-Z.) who performed quality assessment as three independent couples of investigators; the process performed manually and the three couples were blinded to each other’s decisions. In case of disagreement within a couple, D.F, who closely observed the process, was responsible to dissolve any discordance.

### Data synthesis

Data synthesis was performed using MedCalc® Statistical Software version 19.6 (MedCalc Software Ltd, Ostend, Belgium; https://www.medcalc.org; 2020). As effect estimates, mortality rates were extracted from each study and combined together using the random effects, generic inverse variance method of DerSimonian and Laird, which assigned the weight of each study in the pooled analysis inversely to its variance [21]. Random-effects model allows generalizing common effect size beyond the (narrowly defined) population included in the analysis [22]. However, as I^2^ has a substantial bias when the number of studies is small (positive when the true fraction of heterogeneity is small and negative when the true fraction of heterogeneity is large), the point estimate I^2^ should be interpreted cautiously when a meta-analysis has few studies; in fact, in small meta-analyses, confidence intervals should supplement or replace the biased point estimate I^2^ [23].

### Statistical analysis

Analysis of publication bias (small size effect) was performed by funnel plot visualization for asymmetry and use of Egger’s and Begg’s tests. Heterogeneity was based on Q test and I^2^; Q test P value <0.10 and/or I^2^ >50% was indicative of significant heterogeneity and was further analyzed. Analysis of heterogeneity was performed through sensitivity analysis focusing on types of studies, types of acute metabolic emergencies, quality assessment, and GRADE level of evidence to seek whether qualitative or quantitative interaction exists. Univariate comparisons were performed with the use of Pearson’s Chi-square test for discrete variables. All statistical tests were carried out using IBM SPSS Statistics software, version 26.0.0.0, for Windows (IBM Corp ©).

## Results

### Study characteristics

During the final pre-run search prior to the final analysis carried out on January 7, 2021, 313 potentially relevant publications were identified through a thorough search of literature; 174 in EMBASE, 138 in PubMed/Medline, while two more publications of interest were indentified through Google Scholar. A single source of unpublished data of interest was detected. No personal contact with any author that considered necessary contributed any additional information.

After the exclusion of 141 duplicates, all the remaining 173 publications were initially reviewed based only on title and abstract; 110 excluded as being ineligible. Thus, 63 full-text publications were further assessed for eligibility; from these, 39 failed to fulfill the eligibility criteria. The remaining 4 publications included in qualitative synthesis and quantitative synthesis/statistical analysis. These publications included 152 patients (Figure 1).

**Figure 1.**
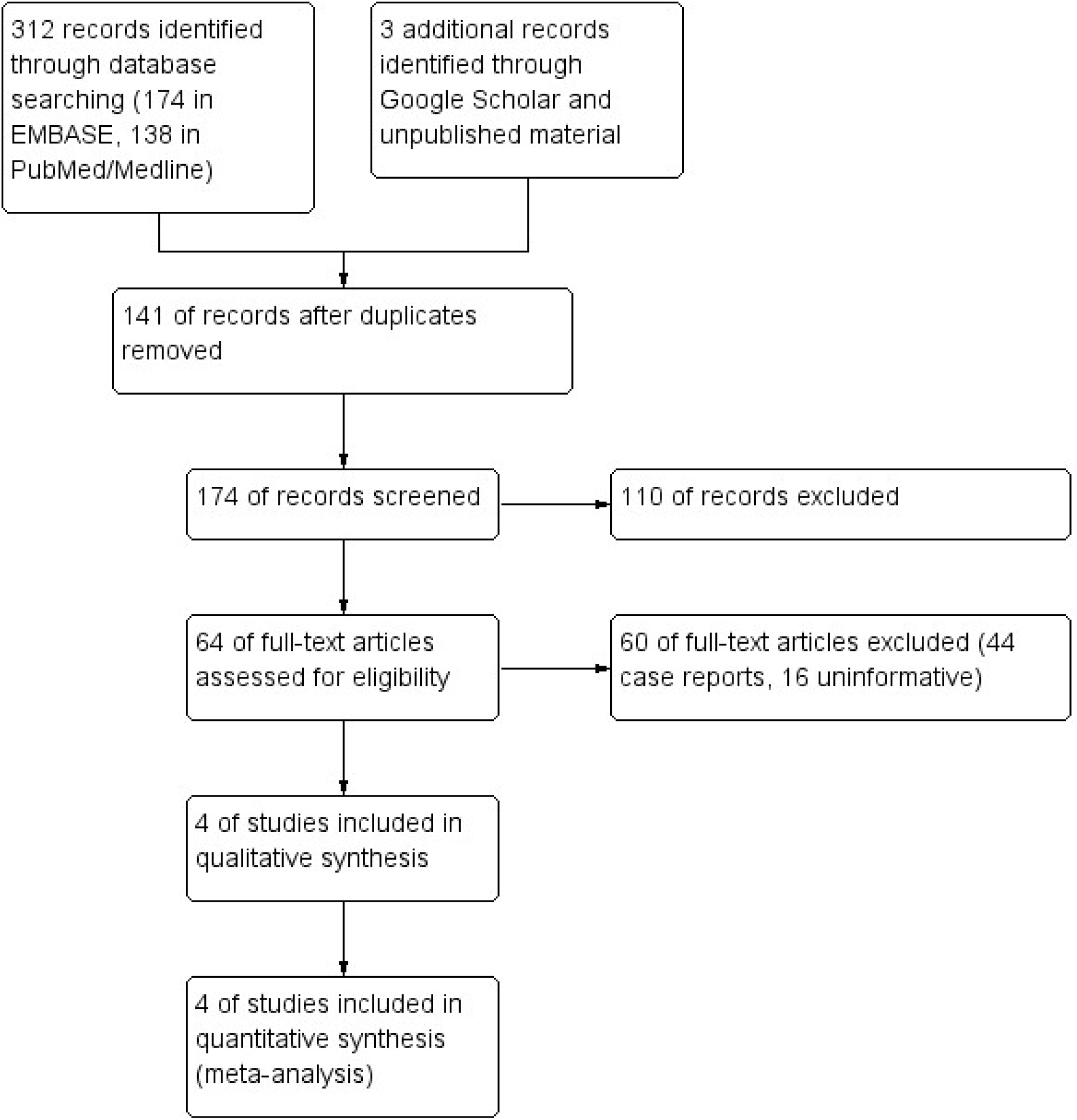
Flow chart

All characteristics regarding title of the study, name of the first author, country where the study was conducted, type of diabetes-related acute metabolic emergency, total number of survivors, total number of non-survivors, and mortality rate were analytically presented in Table 1.

**Table 1.**
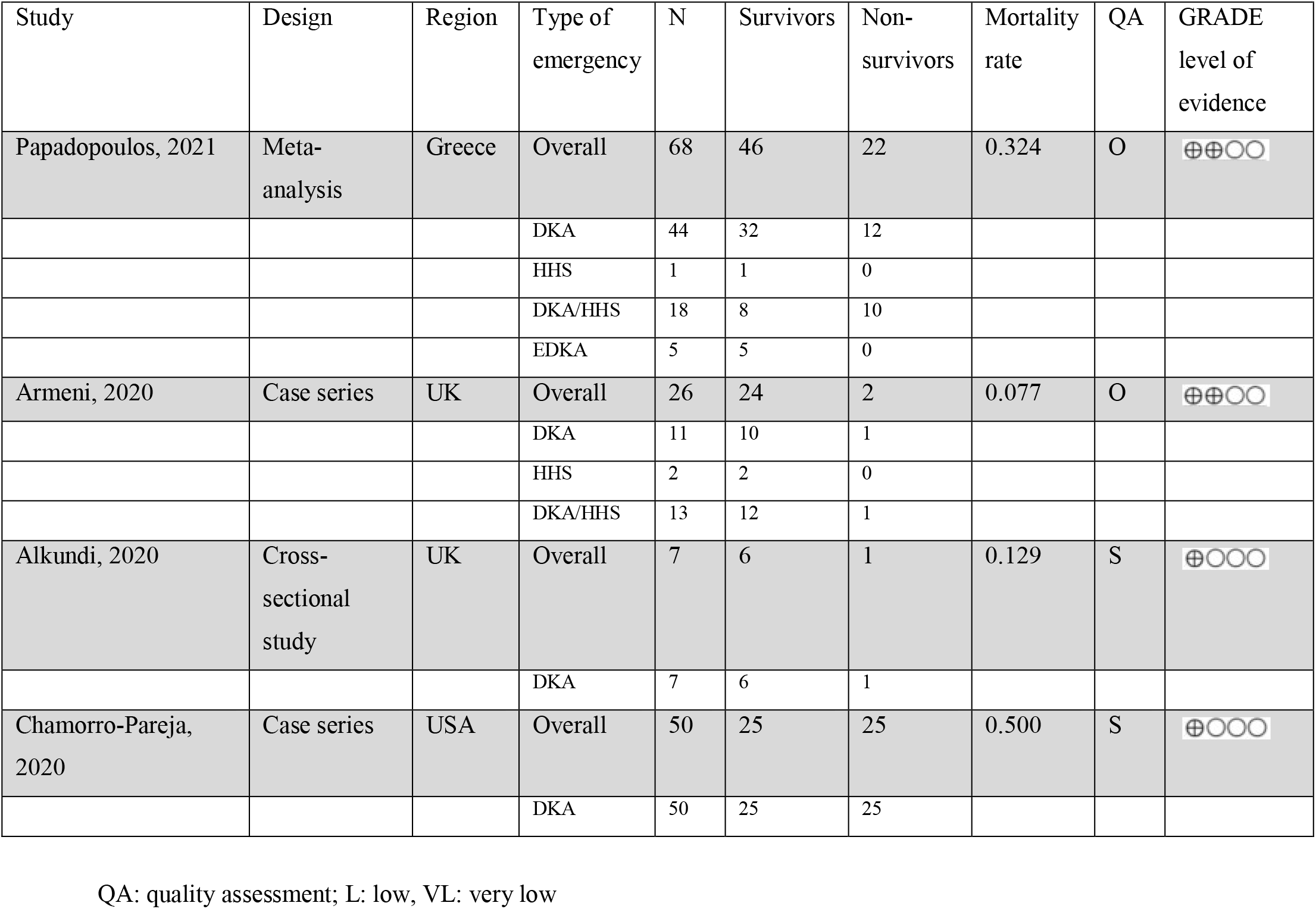
Eligible studies along with study characteristics and quality / risk of bias assessment based on NOS

### Quality assessment and Risk of bias

Quality remarks are provided in Table 1; all details concerning quality assessment items as well as GRADE level of evidence are depicted analytically in Table 2 and Table 3, respectively.

**Table 2.** Quality assessment items of JBI critical appraisal list for case reports concerning included studies (n=2).

| Study | Q1 | Q2 | Q3 | Q4 | Q5 | Q6 | Q7 | Q8 | Optimal quality |
| --- | --- | --- | --- | --- | --- | --- | --- | --- | --- |
| Armeni, 2020 | Yes | Yes | Yes | Yes | Yes | Yes | No | Yes | Yes |
| Chamorro-Pareja, 2020 | Yes | Yes | Yes | Yes | No | No | No | Yes | No |

Q1: Were patient's demographic characteristics clearly described?
Q2: Was the patient's history clearly described and presented as a timeline?
Q3: Was the current clinical condition of the patient on presentation clearly described?
Q4: Were diagnostic tests or assessment methods and the results clearly described?
Q5: Was the intervention(s) or treatment procedure(s) clearly described?
Q6: Was the post-intervention clinical condition clearly described?
Q7: Were adverse events (harms) or unanticipated events identified and described?
Q8: Does the case report provide takeaway lessons?

**Table 3.** Quality assessment items of JBI critical appraisal list for case control studies concerning included studies (n=1); N/A: not applicable; N/R: not reported.

| Study | Q1 | Q2 | Q3 | Q4 | Q5 | Q6 | Q7 | Q8 | Q9 | Q10 | Q11 | Q12 | Optimal quality |
| --- | --- | --- | --- | --- | --- | --- | --- | --- | --- | --- | --- | --- | --- |
| Alkundi, 2020 | Yes | Yes | No | Yes | Yes | Yes | N/A | N/R | Yes | Yes | N/R | No | Fair |

Q1: Was the research question or objective in this paper clearly stated and appropriate?
Q2: Was the study population clearly specified and defined?
Q3: Did the authors include a sample size justification?
Q4: Were controls selected or recruited from the same or similar population that gave rise to the cases (including the same timeframe)?
Q5: Were the definitions, inclusion and exclusion criteria, algorithms or processes used to identify or select cases and controls valid, reliable, and implemented consistently across all study participants?
Q6: Were the cases clearly defined and differentiated from controls?
Q7: If less than 100 percent of eligible cases and/or controls were selected for the study, were the cases and/or controls randomly selected from those eligible?
Q8: Was there use of concurrent controls?
Q9: Were the investigators able to confirm that the exposure/risk occurred prior to the development of the condition or event that defined a participant as a case?
Q10: Were the measures of exposure/risk clearly defined, valid, reliable, and implemented consistently (including the same time period) across all study participants?
Q11: Were the assessors of exposure/risk blinded to the case or control status of participants?
Q12: Were key potential confounding variables measured and adjusted statistically in the analyses? If matching was used, did the investigators account for matching during study analysis?

**Table 4.** GRADE level of evidence concerning included studies (n=4).

| Study | Risk of bias | Imprecision | Inconsistency | Indirectness | Publication bias | Large magnitude of effect | GRADE level of evidence |
| --- | --- | --- | --- | --- | --- | --- | --- |
| Papadopoulos, 2021 | No serious risk of bias | No serious imprecision | No serious inconsistency | No serious indirectness | No serious publication bias | No | Low |
| Armeni, 2020 | No serious risk of bias | No serious imprecision | N/A | No serious indirectness | N/A | No | Low |
| Alkundi, 2020 | Serious | No serious imprecision | N/A | No serious indirectness | N/A | No | Very Low |
| Chamorro-Pareja, 2020 | Serious | No serious imprecision | N/A | No serious indirectness | N/A | No | Very Low |

N/A: Not applicable.

**Table 5.** Sensitivity analysis.

|  | Mortality rate |  | I<sup>2</sup> |  |  |
| --- | --- | --- | --- | --- | --- |
|  | Combined mean | 95% CI | Mean | 95% CI | P |
| Overall result |  |  |  |  |  |
| All studies included | 0.271 | 0.112-0.469 | 82.8 | 56.1-93.3 | 0.0006 |
| Sensitivity analysis according to study type |  |  |  |  |  |
| Meta-analyses excluded | 0.241 | 0.027-0.574 | 88.6 | 68.4-95.9 | 0.0002 |
| Sensitivity analysis according to emergency type |  |  |  |  |  |
| Only DKA patients included | 0.289 | 0.134-0.475 | 71.8 | 20.0-90.1 | 0.0138 |
| Only HHS patients included | 0.112 | 0.009-0.495 | 0.0 | 0.0-0.0 | 0.8523 |
| Only EDKA patients included | 0.000 | N/A | N/A | N/A | N/A |
| Only DKA/HHS patients included | 0.315 | 0.012-0.780 | 86.8 | 48.2-96.7 | 0.0058 |
| Sensitivity analysis according to quality assessment |  |  |  |  |  |
| Studies of “suboptimal” quality excluded | 0.203 | 0.030-0.475 | 85.7 | 42.5-96.4 | 0.0082 |
| Sensitivity analysis according to GRADE level of evidence |  |  |  |  |  |
| Studies of GRADE “very low” level of evidence excluded | 0.203 | 0.030-0.475 | 85.7 | 42.5-96.4 | 0.0082 |
| Sensitivity analysis at study level |  |  |  |  |  |
| Papadopoulos, 2021 excluded | 0.241 | 0.027-0.574 | 88.6 | 68.4-95.9 | 0.0002 |
| Armeni, 2020 excluded | 0.360 | 0.207-0.529 | 67.2 | 0.0-90.5 | 0.0472 |
| Alkundi, 2020 excluded | 0.296 | 0.110-0.528 | 87.6 | 65.1-95.6 | 0.0003 |
| Chamorro-Pareja, 2020 excluded | 0.196 | 0.057-0.391 | 72.9 | 8.7-91.9 | 0.0251 |

N/A: Not applicable.

### Primary outcome

Combined mortality rate was found to be 27.1% [95% CI: 11.2-46.9%] (Figure 2). Heterogeneity was considerable (I^2^=83%; 95% CI: 56-93%), corrected to 67% according to Von Hippel adjustment for small meta-analyses; this value was based on an approximation for I^2^=80% yielding a real value 64%, and consequently, an bias leading to 16% overestimation (Supplementary Figure 1).

**Figure 2.**
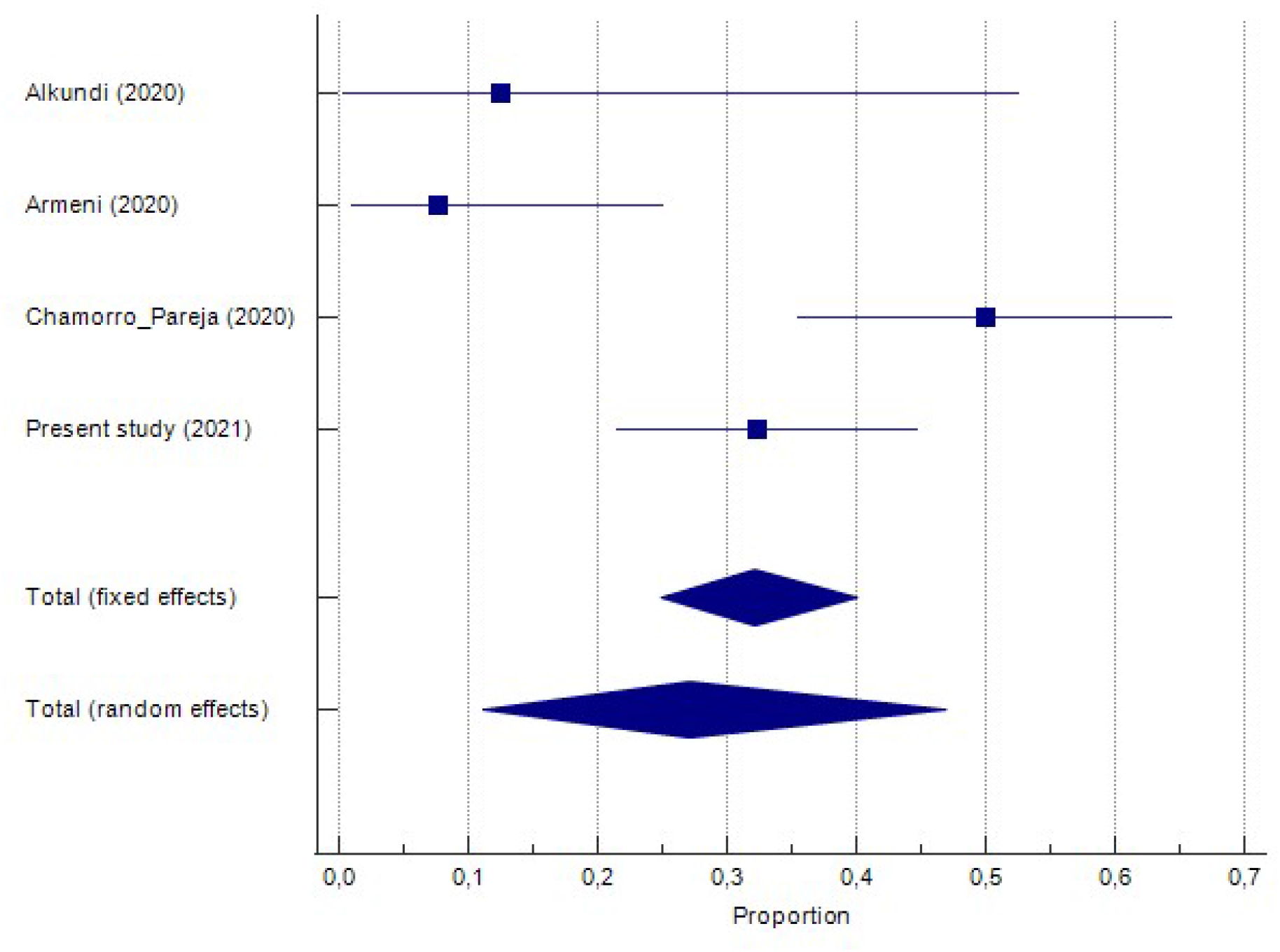
Forest plot depicting combined mortality rate from diabetes-associated acute metabolic emergencies in COVID-19 patients.

**Figure 3.**
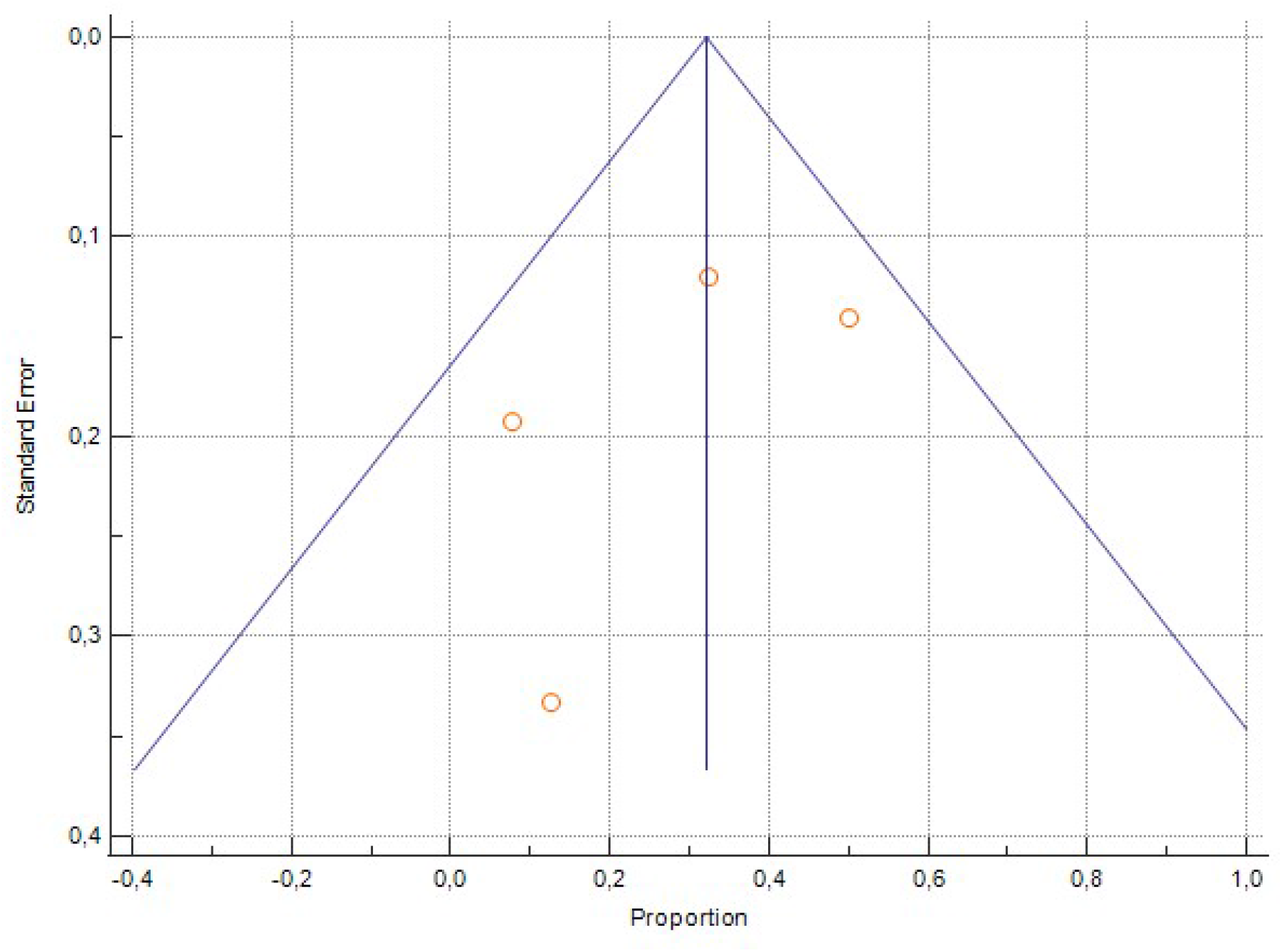
Funnel plot showing no apparent asymmetry.

### Publication bias (small size effect)

No significant publication bias (small size effect) was detected as funnel plot presented no apparent asymmetry. Moreover, both Egger’s and Begg’s tests yielded an insignificant result (P=0.44 and P=0.50, respectively).

### Analysis of heterogeneity

Sensitivity analysis was carried out according to: i) study type (meta-analyses included vs excluded), ii) emergency type (DKA patients included vs excluded, EDKA patients included vs excluded, HHS patients included vs excluded, DKA/HHS patients included vs excluded), iii) quality assessment (studies of “suboptimal” quality included vs excluded), iv) GRADE level of evidence (studies of “very low” level of evidence excluded vs included); furthermore, sensitivity analysis was performed at single study level. There was no difference as deduced by the inspection of the relevant confidence intervals and thus, sensitivity analysis failed to explain the observed heterogeneity.

## Discussion

In the present study we have demonstrated that diabetes-associated acute metabolic emergencies in COVID-19 patients (DKA, HHS, EDKA, and DKA/HHS) are characterized by considerable mortality, which has been estimated to be 27.1% [95% CI: 11.2-46.9%].

Our findings are in keeping with a recent meta-analysis of 41 case reports carried out by our team, which yielded a mortality rate of 32.4% among 68 patients with known outcome [13]. We are totally aware that including a self-report in a meta-analysis can import a severe bias. However, there are at least four reasons which alleviate this danger: first, the meta-analysis of Papadopoulos et al. exhibits the least deviation from the vertical line of the funnel plot, representing the mean; second, sensitivity analysis did not reveal any profound difference regarding combined mortality rate and I^2^; third, it is of “optimal” quality and has a GRADE “low” level of evidence, namely the best that a study of such a kind can achieve at first; fourth, it is the most representative as it is the only that includes patients presenting all four different kind of emergencies (DKA, HHS, EDKA, and combined DKA/HHS) as well as patients from 41 different sources.

Armeni et al. had a substantial contribution to the topic by analytically describing 35 patients with COVID-19, 26 of which had either DKA (n=11), or HHS (n=2), or mixed DKA/HHS (n=13). This study, although being a case series, is multicenter and of well established quality [8].

Interestingly, Chamorro-Pareja et al. reported an unusually high mortality rate among 50 COVID-19 patients who presented DKA (50%) [11]. In fact, comparing relevant data from all included studies over a 2×4 contingency table (df=3), a statistically significant result is obtained (Chi-square P=0.014); this variability is partly explained by the meta-analysis of Papadopoulos et al. who reported that, apart from the presence of mixed DKA/HHS, two other parameters, namely the presence of acute kidney injury as well as the necessity for mechanical ventilation of COVID-19 patients (critical illness or disease status 4 illness) are key determinants of outcome [13].

Alkundi et al. report the very intriguing - if not controversial - finding that COVID-19 patients presented with DKA, when compared with COVID-19 patients who had not developed DKA, were more likely to survive (P=0.046). Their analysis was carried out with the use of Kaplan-Meier survival curves. However, the authors did not adjust their finding for potent confounders, using Cox-regression, most probably due to the small number of sample size. As a matter of fact, their result needs at least to be considered cautiously and has to be further evaluated in larger studies [12].

The major limitation of the present study might be dual: first, the combination of data from different kind of studies, namely two case report series, one case-control study, and one meta-analysis of 41 case reports; second, the very small number of studies included. However, as the topic is totally novel, any study that respects adherence to protocol followed, investigates causes of heterogeneity, and assesses the impact of risk of bias on the evidence synthesis might be valuable [24].

A serious query could focus on the decision to proceed to the meta-analysis despite the considerable amount of heterogeneity. However, several reasons might support our approach: 1) there was little evidence of publication bias (as funnel plot did not decline from asymmetry), there was no evidence of small size studies effect (as Egger’s and Begg’s tests were not statistically significant), 3) there was no considerable qualitative interaction.

In conclusion, the present meta-analysis illustrated that COVID-19 related acute metabolic emergencies (DKA, HHS, and EDKA) are characterized by considerable mortality; thus, clinicians should be aware of timely detection and immediate treatment commencing. Future, cumulative evidence are welcome to further enlighten this field.

## Data Availability

All data referred to the manuscript are totally available.

## Compliance with Ethical Standards

### Research involving Human Participants and/or Animals

All procedures performed in studies involving human participants were in accordance with the ethical standards of the institutional and/or national research committee and with the 1964 Helsinki declaration and its later amendments or comparable ethical standards.

This article does not contain any studies with animals performed by any of the authors.

### Informed consent

Informed consent was obtained from all individual participants included in the study.

## Figure legends

**Supplementary Figure 1.**
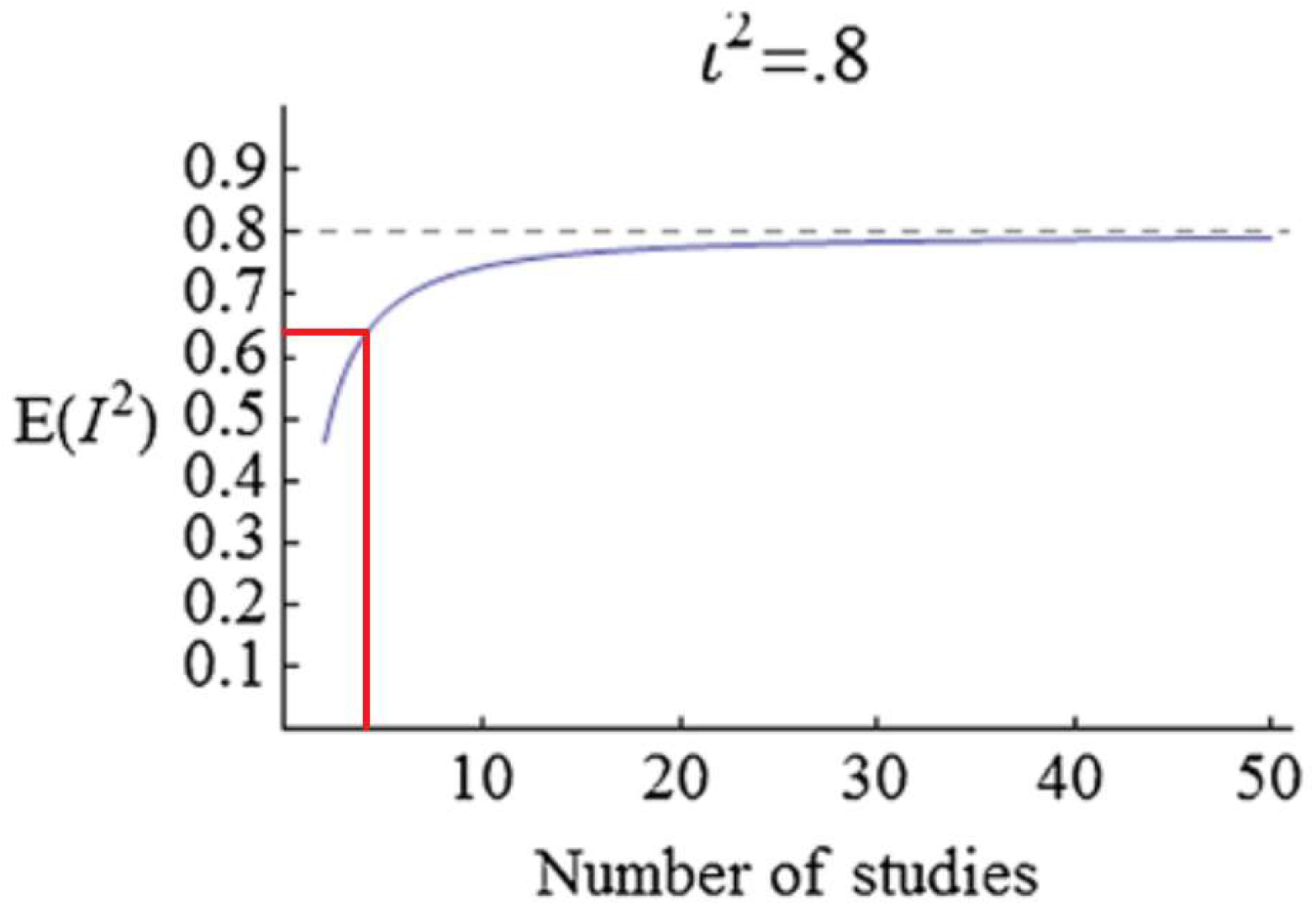
Von Hippel adjustment for small meta-analyses corrected observed heterogeneity (I^2^) corrects 80% to ∼64% in case that the number of included studies is n = 4; consequently, I^2^=83 is estimated to be corrected to ∼67%.

## References

1. Zhou F, Yu T, Du R, Fan G, Liu Y, Liu Z, Xiang J, Wang Y, Song B, Gu X, Guan L, Wei Y, Li H, Wu X, Xu J, Tu S, Zhang Y, Chen H, Cao B. Clinical course and risk factors for mortality of adult inpatients with COVID-19 in Wuhan, China: a retrospective cohort study. Lancet. 2020 Mar 28;395(10229):1054-1062. doi: 10.1016/S0140-6736(20)30566-3. Epub 2020 Mar 11. Erratum in: Lancet. 2020 Mar 28;395(10229):1038. Erratum in: Lancet. 2020 Mar 28;395(10229):1038. PMID: 32171076; PMCID: PMC7270627.

2. Gregory JM, Slaughter JC, Duffus SH, Smith TJ, LeStourgeon LM, Jaser SS, McCoy AB, Luther JM, Giovannetti ER, Boeder S, Pettus JH, Moore DJ. COVID-19 Severity Is Tripled in the Diabetes Community: A Prospective Analysis of the Pandemic’s Impact in Type 1 and Type 2 Diabetes. Diabetes Care. 2020 Dec 2:dc202260. doi: 10.2337/dc20-2260. Epub ahead of print. PMID: 33268335.

3. McGurnaghan SJ, Weir A, Bishop J, Kennedy S, Blackbourn LAK, McAllister DA, Hutchinson S, Caparrotta TM, Mellor J, Jeyam A, O’Reilly JE, Wild SH, Hatam S, Höhn A, Colombo M, Robertson C, Lone N, Murray J, Butterly E, Petrie J, Kennon B, McCrimmon R, Lindsay R, Pearson E, Sattar N, McKnight J, Philip S, Collier A, McMenamin J, Smith-Palmer A, Goldberg D, McKeigue PM, Colhoun HM; Public Health Scotland COVID-19 Health Protection Study Group; Scottish Diabetes Research Network Epidemiology Group. Risks of and risk factors for COVID-19 disease in people with diabetes: a cohort study of the total population of Scotland. Lancet Diabetes Endocrinol. 2020 Dec 23:S2213-8587(20)30405-8. doi: 10.1016/S2213-8587(20)30405-8. Epub ahead of print. PMID: 33357491.

4. Al Hayek AA, Robert AA, Alotaibi ZK, Al Dawish M. Clinical characteristics of hospitalized and home isolated COVID-19 patients with type 1 diabetes. Diabetes Metab Syndr. 2020 Sep 10;14(6):1841–1845. doi: 10.1016/j.dsx.2020.09.013. Epub ahead of print. PMID: 32971511; PMCID: PMC7481862.

5. Chowdhury S, Goswami S. COVID-19 and type 1 diabetes: dealing with the difficult duo. Int J Diabetes Dev Ctries. 2020 Jul 14:1–6. doi: 10.1007/s13410-020-00846-z. Epub ahead of print. PMID: 32837091; PMCID: PMC7359765.

6. Wu C, Chen X, Cai Y, Xia J, Zhou X, Xu S, Huang H, Zhang L, Zhou X, Du C, Zhang Y, Song J, Wang S, Chao Y, Yang Z, Xu J, Zhou X, Chen D, Xiong W, Xu L, Zhou F, Jiang J, Bai C, Zheng J, Song Y. Risk Factors Associated With Acute Respiratory Distress Syndrome and Death in Patients With Coronavirus Disease 2019 Pneumonia in Wuhan, China. JAMA Intern Med. 2020 Jul 1;180(7):934–943. doi: 10.1001/jamainternmed.2020.0994. PMID: 32167524; PMCID: PMC7070509.

7. Yang JK, Lin SS, Ji XJ, Guo LM. Binding of SARS coronavirus to its receptor damages islets and causes acute diabetes. Acta Diabetol. 2010 Sep;47(3):193–9. doi: 10.1007/s00592-009-0109-4. Epub 2009 Mar 31. PMID: 19333547; PMCID: PMC7088164.

8. Armeni E, Aziz U, Qamar S, Nasir S, Nethaji C, Negus R, Murch N, Beynon HC, Bouloux P, Rosenthal M, Khan S, Yousseif A, Menon R, Karra E. Protracted ketonaemia in hyperglycaemic emergencies in COVID-19: a retrospective case series. Lancet Diabetes Endocrinol. 2020 Aug;8(8):660–663. doi: 10.1016/S2213-8587(20)30221-7. Epub 2020 Jul 1. PMID: 32621809; PMCID: PMC7329282.

9. Tsai PH, Lai WY, Lin YY, Luo YH, Lin YT, Chen HK, Chen YM, Lai YC, Kuo LC, Chen SD, Chang KJ, Liu CH, Chang SC, Wang FD, Yang YP. Clinical Manifestation and Disease Progression in COVID-19 Infection. J Chin Med Assoc. 2020 Nov 19;Publish Ahead of Print. doi: 10.1097/JCMA.0000000000000463. Epub ahead of print. PMID: 33230062.

10. Muniangi-Muhitu H, Akalestou E, Salem V, Misra S, Oliver NS, Rutter GA. Covid-19 and Diabetes: A Complex Bidirectional Relationship. Front Endocrinol (Lausanne). 2020 Oct 8;11:582936. doi: 10.3389/fendo.2020.582936. PMID: 33133024; PMCID: PMC7578412.

11. Chamorro-Pareja N, Parthasarathy S, Annam J, Hoffman J, Coyle C, Kishore P. Letter to the editor: Unexpected high mortality in COVID-19 and diabetic ketoacidosis. Metabolism. 2020 Sep;110:154301. doi: 10.1016/j.metabol.2020.154301. Epub 2020 Jun 24. PMID: 32589899; PMCID: PMC7311346.

12. Alkundi A, Mahmoud I, Musa A, Naveed S, Alshawwaf M. Clinical characteristics and outcomes of COVID-19 hospitalized patients with diabetes in the United Kingdom: A retrospective single centre study. Diabetes Res Clin Pract. 2020 Jul;165:108263. doi: 10.1016/j.diabres.2020.108263. Epub 2020 Jun 10. PMID: 32531325; PMCID: PMC7283049.

13. Papadopoulos VP, Koutroulos MV, Zikoudi DG, Bakola SA, Avramidou P, Touzlatzi N, Filippou DK. Acute Metabolic Emergencies in Diabetes and COVID-19: a systematic review and meta-analysis of case reports. MedRxiv 2021.01.10.21249550; doi: https://doi.org/10.1101/2021.01.10.21249550

14. Stroup DF, Berlin JA, Morton SC, Olkin I, Williamson GD, Rennie D, Moher D, Becker BJ, Sipe TA, Thacker SB. Meta-analysis of observational studies in epidemiology: a proposal for reporting. Meta-analysis Of Observational Studies in Epidemiology (MOOSE) group. JAMA. 2000 Apr 19;283(15):2008–12. doi: 10.1001/jama.283.15.2008. PMID: 10789670.

15. Shea BJ, Reeves BC, Wells G, Thuku M, Hamel C, Moran J, Moher D, Tugwell P, Welch V, Kristjansson E, Henry DA. AMSTAR 2: a critical appraisal tool for systematic reviews that include randomised or non-randomised studies of healthcare interventions, or both. BMJ. 2017 Sep 21;358:j4008. doi: 10.1136/bmj.j4008. PMID: 28935701; PMCID: PMC5833365.

16. Ma LL, Wang YY, Yang ZH, Huang D, Weng H, Zeng XT. (2020) Methodological quality (risk of bias) assessment tools for primary and secondary medical studies: what are they and which is better? Mil Med Res. 7(1):7. doi:10.1186/s40779-020-00238-8.

17. Moola S, Munn Z, Tufanaru C, Aromataris E, Sears K, Sfetcu R, Currie M, Lisy K, Qureshi R, Mattis P, Mu P. Chapter 7: Systematic reviews of etiology and risk. In: Aromataris E, Munn Z (Editors). JBI Manual for Evidence Synthesis. JBI, 2020. Available from https://synthesismanual.jbi.global, https://doi.org/10.46658/JBIMES-20-08.

18. Guyatt GH, Oxman AD, Kunz R, Vist GE, Falck-Ytter Y, Schünemann HJ; GRADE Working Group. What is “quality of evidence” and why is it important to clinicians? BMJ. 2008 May 3;336(7651):995–8. doi: 10.1136/bmj.39490.551019.BE. PMID: 18456631; PMCID: PMC2364804.

19. Guyatt GH, Oxman AD, Vist GE, Kunz R, Falck-Ytter Y, Alonso-Coello P, Schünemann HJ; GRADE Working Group. GRADE: an emerging consensus on rating quality of evidence and strength of recommendations. BMJ. 2008 Apr 26;336(7650):924–6. doi: 10.1136/bmj.39489.470347.AD. PMID: 18436948; PMCID: PMC2335261.

20. Guyatt G, Oxman AD, Akl EA, Kunz R, Vist G, Brozek J, Norris S, Falck-Ytter Y, Glasziou P, DeBeer H, Jaeschke R, Rind D, Meerpohl J, Dahm P, Schünemann HJ. GRADE guidelines: 1. Introduction-GRADE evidence profiles and summary of findings tables. J Clin Epidemiol. 2011 Apr;64(4):383–94. doi: 10.1016/j.jclinepi.2010.04.026. Epub 2010 Dec 31. PMID: 21195583.

21. DerSimonian R, Laird N. (2015) Meta-analysis in clinical trials revisited. Contemp Clin Trials. 45(Pt A):139–145. doi:10.1016/j.cct.2015.09.002.

22. Borenstein M, Hedges LV, Higgins JP, Rothstein HR. (2010) A basic introduction to fixed-effect and random-effects models for meta-analysis. Res Synth Methods. 1(2):97–111. doi:10.1002/jrsm.12.

23. Von Hippel PT. The heterogeneity statistic I^2^ can be biased in small meta-analyses. BMC Med Res Methodol. 2015 Apr 14;15:35. doi: 10.1186/s12874-015-0024-z. PMID: 25880989; PMCID: PMC4410499.

24. Rainkie DC, Abedini ZS, Abdelkader NN. Reporting and methodological quality of systematic reviews and meta-analysis with protocols in Diabetes Mellitus Type II: A systematic review. PLoS One. 2020 Dec 16;15(12):e0243091. doi: 10.1371/journal.pone.0243091. PMID: 33326429; PMCID: PMC7743973.

